# Global Perspective of COVID-19 Vaccine Nationalism

**DOI:** 10.1101/2021.12.31.21268580

**Authors:** Palash Basak, Tanvir Abir, Abdullah Al Mamun, Noor Raihani Zainol, Mansura Khanam, Md. Rashidul Haque, Abul Hasnat Milton, Kingsley Emwinyore Agho

## Abstract

This study aimed to explore the global perspective of the association between GDP of various countries and progress of COVID-19 vaccinations; to explore how the global pattern holds in the continents, and investigate the spatial distribution pattern of COVID-19 vaccination progress for all countries. We have used consolidated data on COVID-19 vaccination and GDP from Our World in Data, an open-access data source. Data analysis and visualization were performed in R-Studio. There was a strong linear association between per capita income and the proportion of people vaccinated in countries with one million or more populations. GDP per capita accounts for a 50% variation in the vaccination rate across the nations. Our assessments revealed that the global pattern holds in every continent. Rich European and North-American countries are most protected against COVID-19. Less developed African countries barely initiated the vaccination program. There is a significant disparity among Asian countries. The security of wealthier nations (vaccinated their citizens) cannot be guaranteed unless adequate vaccination covers the less-endowed countries. Therefore, the global community should take initiatives to speed up the COVID-19 vaccination program in all countries of the world, irrespective of their wealth.

## 1. Introduction

The worldwide effort to work up safe and effective Covid-19 vaccines has produced remarkable results, thanks in part to early, crucial investments in clinical discovery through initiatives like Operation Warp Speed (Kim, Hotez et al., 2021). These accomplishments demonstrate the benefits of consistent, extended funding for basic research and immunology: the scientific community was prepared to take action. Now, as the world is faced with a scarcity of vaccines, there is a depressing reality: As of December 25, 2021, 57.4% of the world population has received at least one dose of a COVID-19 vaccine (Ritchie, Mathieu et al. 2020); approximately 8.95 billion doses have been administered globally, 37.19 million are now administered each day (Ritchie, Mathieu et al. 2020). However, only 8.3% of people from low-income countries have received at least one dose (Ritchie, Mathieu et al. 2020). Vaccine distribution is still very low in many of the world’s poorest countries. Experts predicted that 20% of the population in low-resource areas would receive a vaccine in 2021 (Katz, Weintraub et al. 2021), but in reality, the actual achievement was much lower. The critical nature of investment in research notwithstanding, prolonged neglect of public health and global delivery strategies has rendered humankind unprepared to bring this pandemic to an end. Priority must be given to solving the complex bottlenecks in distributing and allocating newly approved vaccines (Katz, Weintraub et al., 2021). As part of these efforts, vaccines must be produced in a safe, efficient, and timely manner. As a result of mistrust, misinformation, and historical legacies, vaccine adoption is hindered (Weintraub, Subramanian et al., 2021). Even wealthy countries have encountered formidable obstacles when implementing mass vaccination programs and made critical mistakes (Katz, Weintraub et al., 2021).

Aside from that, the early competitive procurement of vaccines by the United States and purchases by other high-income countries has led to the universal assumption that each country will be exclusively responsible for its population. When powerful countries secure vaccines and therapies at the expense of less-wealthy countries, it perpetuates a long history of shortsightedness, inefficiency, and death (Katz, Weintraub et al., 2021). Industrialized countries are anxious to help with global vaccination, particularly for countries that 76 require partnerships to warrant supply and delivery. Uncoordinated patches of immunity could also exacerbate the spread of escape variants (Katz, Weintraub et al., 2021). As a result of these inequalities, vaccines and essential medications are treated as a market commodity instead of a public good in global health and more widely in our global economy.

Similar policies have been implemented in the past during pandemics. Antiretroviral therapy was out of reach for most low-resource countries when HIV was at its zenith because of prohibitively high prices imposed by the pharmaceutical industry, as well as a belief among United Nations agencies and major donors that prevention should take precedence over treatment (Katz, Weintraub et al., 2021). Inequities in access and health and economic well-being are exacerbated by the commodification of public goods in the global economy. As a moral and national security issue, eliminating critical constraints requires bold, decisive action to ensure the expansion of supply and delivery of Covid-19 vaccines. As part of the Covid-19 Vaccines Global Access (COVAX) program, which supplies vaccines to low- and middle-income countries, the United States, under the Biden administration, and the G7 nations have pledged support for global vaccine procurement. Still, this funding is insufficient (Katz, Weintraub et al. 2021). Currently, COVAX plans to vaccinate at least 20 percent of participating countries’ populations by the year 2021. Even though this would be a significant achievement, it falls far short of the goal of quickly securing global herd immunity (Mullard 2020).

Even prior to any vaccine’s approval, high-income countries that constitute only a fraction of the global population had already placed orders for more than 50% of the projected early supply of doses for COVID-19 vaccines. By mid-August of 2020, the US had secured 800 million doses of at least six different vaccines in development; the UK had procured 340 million doses, with around 5 per capita; and the European Union (EU) and Japan had each placed orders for hundreds of millions of doses (Callaway 2020). While the world’s wealthiest nations have booked enough doses of the best COVID-19 vaccines to immunise their individual populations numerous times (Bhutto 2021), forecast global manufacturing capacity also implies limited and delayed access of low-income nations to this significant healthcare resource. This current study examined the association between national income and COVID-19 vaccination in countries with 1 million or more population. It highlights how vaccine nationalism in the context of COVID-19 has uncovered the risks of nationalist responses to a simultaneous global emergency and how it thus represents a pivotal moment in which the dynamics, courses, and directions of contemporary globalization processes must be crucially reflected on and acted upon.

## 2. Materials and Methods

### 2.1 Data

The Coronavirus Pandemic (COVID-19) dataset used in the research was downloaded from Our World in Data (Ritchie, Mathieu et al. 2020) at 07:43 pm on December 25, 2021, Australian Eastern Daylight Time (AEDT). Along with the vaccination rate, the dataset contained information about GDP. For exploring evidence of vaccine nationalism, both vaccination rate and GDP data were used. Countries with a population of less than one million were excluded from the study to focus on sizable nations. The dataset was also filtered for null values. A total of 153 countries with one million or more population were included in the study, and their data on GDP and vaccination were available.

### 2.2 Analysis

R-Studio Cloud (RStudio Workbench, Version 2021.09.1 Build 372.pro1, https://rstudio.cloud) was used for data gathering and reorganization and statistical analysis. Graphs were also constructed with the same application. Esri’s ArcGIS Pro (version 2.9, esri.com/en-us/arcgis/products/arcgis-pro) was used to prepare the map.

## 3. Results

Figure 1 shows a strong linear association between GDP per capita and the proportion of the people vaccinated (partially or fully) in 153 countries of the world (*r* = .79, *p* < .001), where data for both vaccination rates and GDP were available. GDP per capita (log_10_) explains about 63% variation in the vaccination rate across the countries. In general, the wealthier the nation is, the higher the vaccination rate. The top three countries with the highest vaccination rate are United Arab Emirates (UAE), Portugal, and Chile. GPD per capita of these top-performing countries is more than US$ 22,000, and their vaccination rate is above 89%. In contrast, the bottom three countries are Burundi, the Democratic Republic of Congo (DRC), and Haiti. GPD of these countries is less than US$1,700, and their vaccination rate is 1% or less.

**Figure 1.**
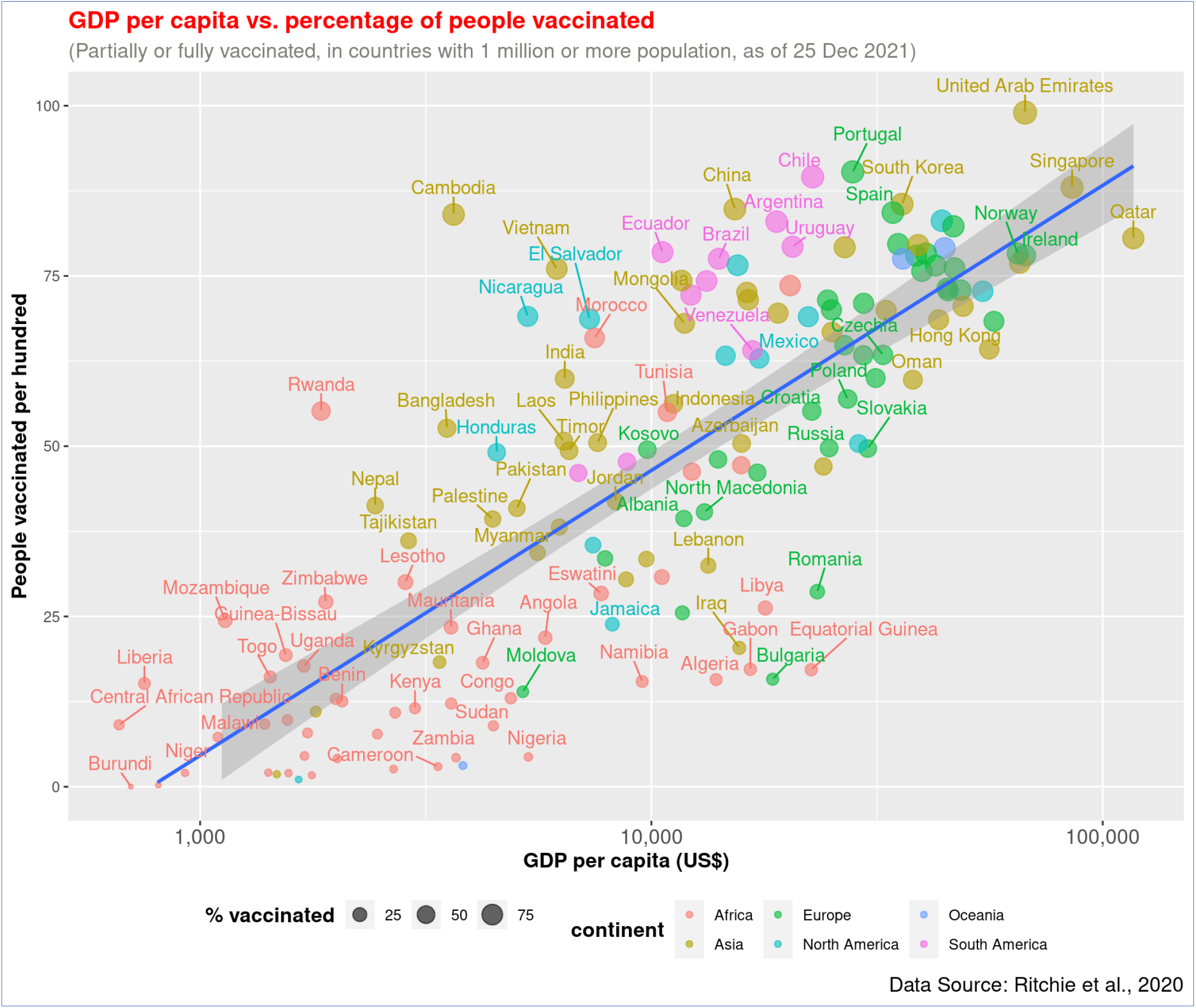
Association of GDP per capita and COVID-19 vaccination rate (full or partial) across countries. The regression line has been presented with blue colour. The figure shows that wealthier nations got higher vaccination rate.

Similarly, a statistically significant correlation was also observed between GDP per capita (log_10_) and full vaccination rate, *r* = .82, *p* < 0.001, there is a linear association between these two variables (Figure 2). Countries like UAE, Portugal, and Singapore have already provided full vaccination to over 87% of people, while Burundi, DRC, and Chad barely immunized people with two doses (less than 0.5%). The data on partial and full vaccination rates are highly correlated, *r* = 0.98, *p* < 0.001 (Figure 3). The countries with higher partial vaccination also achieved higher full vaccination rates.

**Figure 2.**
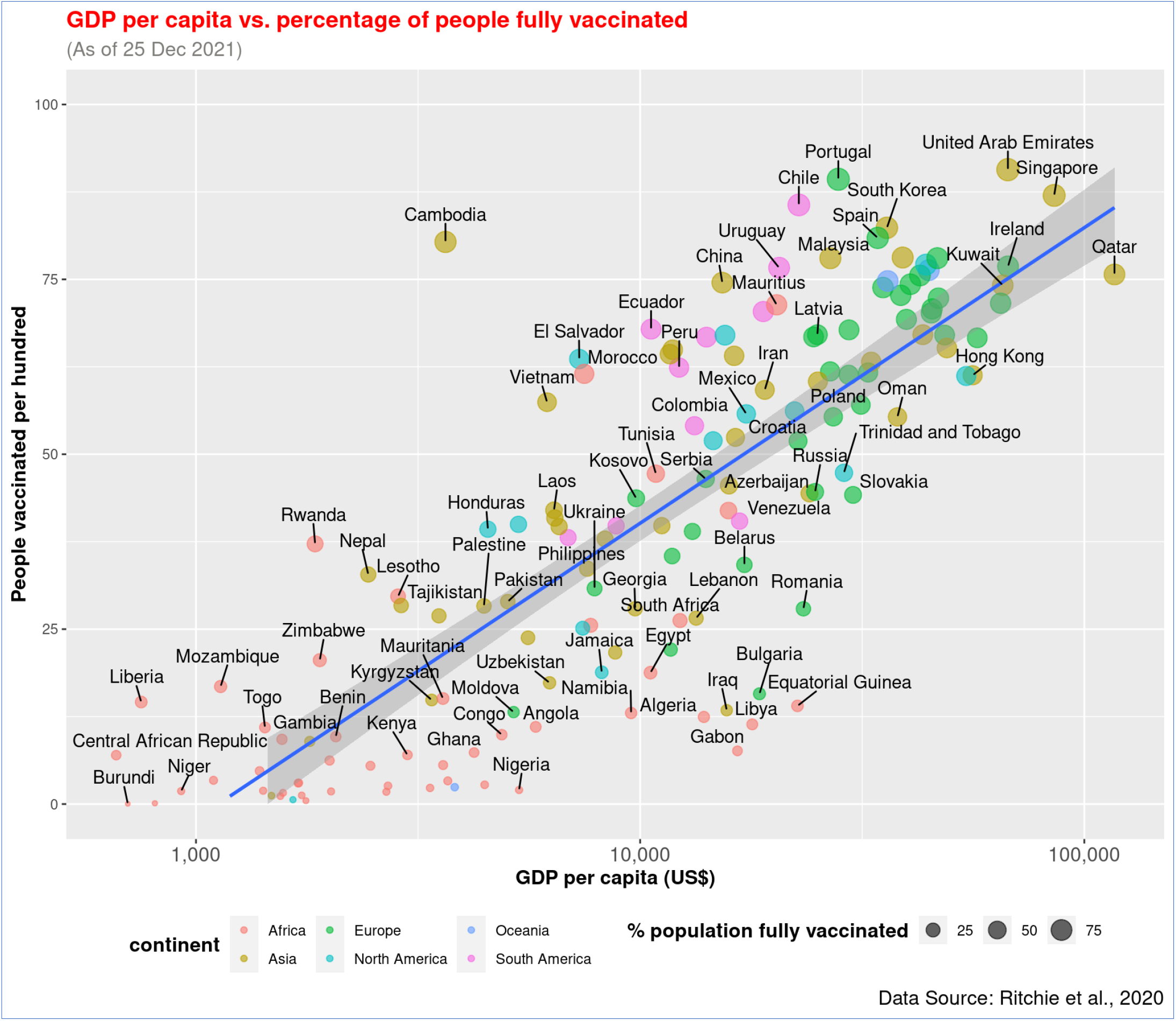
Association of GDP per capita and COVID-19 full vaccination rate across countries. The relationship of both partial and full vaccination rates with GDP is similar.

**Figure 3.**
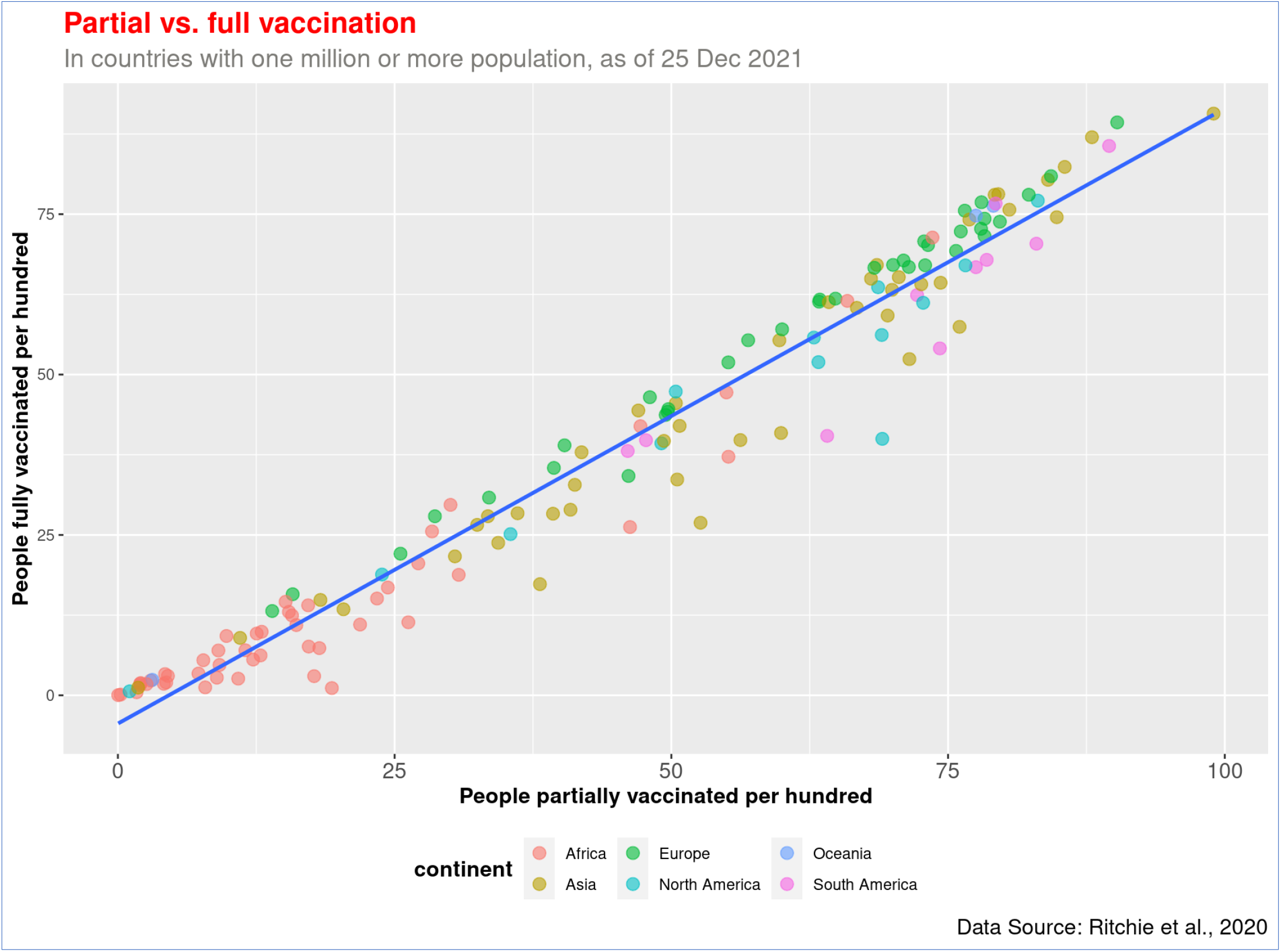
Association between partial and full vaccination. Countries that have achieved a higher level of partial vaccination also reached a higher full vaccination, and most African countries are on the lower left side of the graph.

The global pattern of the association between GDP per capita and vaccination holds across continents (Figures 4 and 5). The richer the country, the higher the vaccination rate is in every continent. The wealthy countries in Europe, North America, and Asia managed to protect a higher proportion of the population with the vaccine. As shown in Figures 4 and 5, African countries have the lowest income. Except few, such as Mauritius and Morocco, all other countries in that continent are lagging in COVID-19 vaccination.

**Figure 4.**
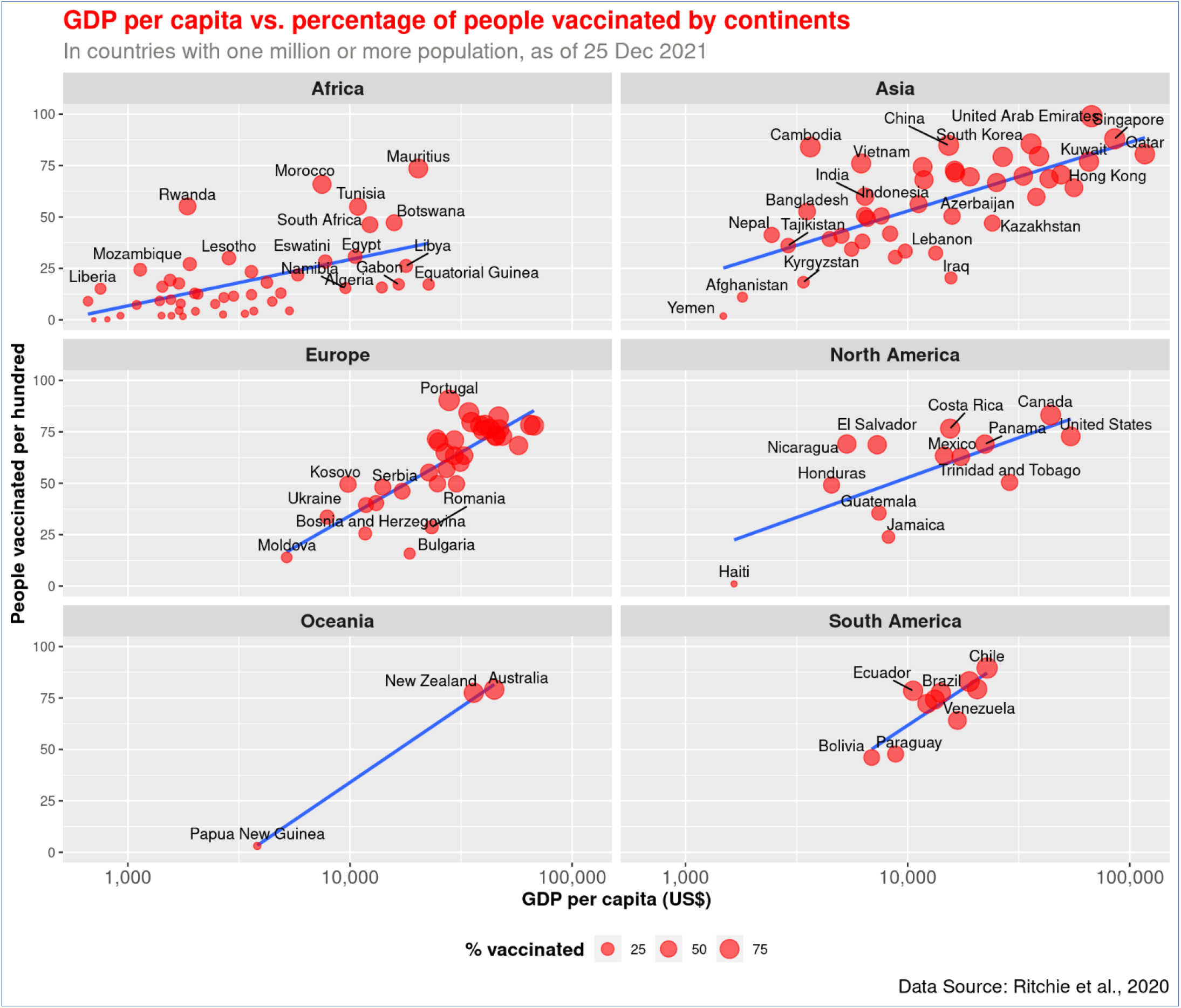
Association of GDP per capita and COVID-19 vaccination rate (full or partial) by continents. The figure shows that wealthier nations got higher vaccination rates on all continents.

**Figure 5.**
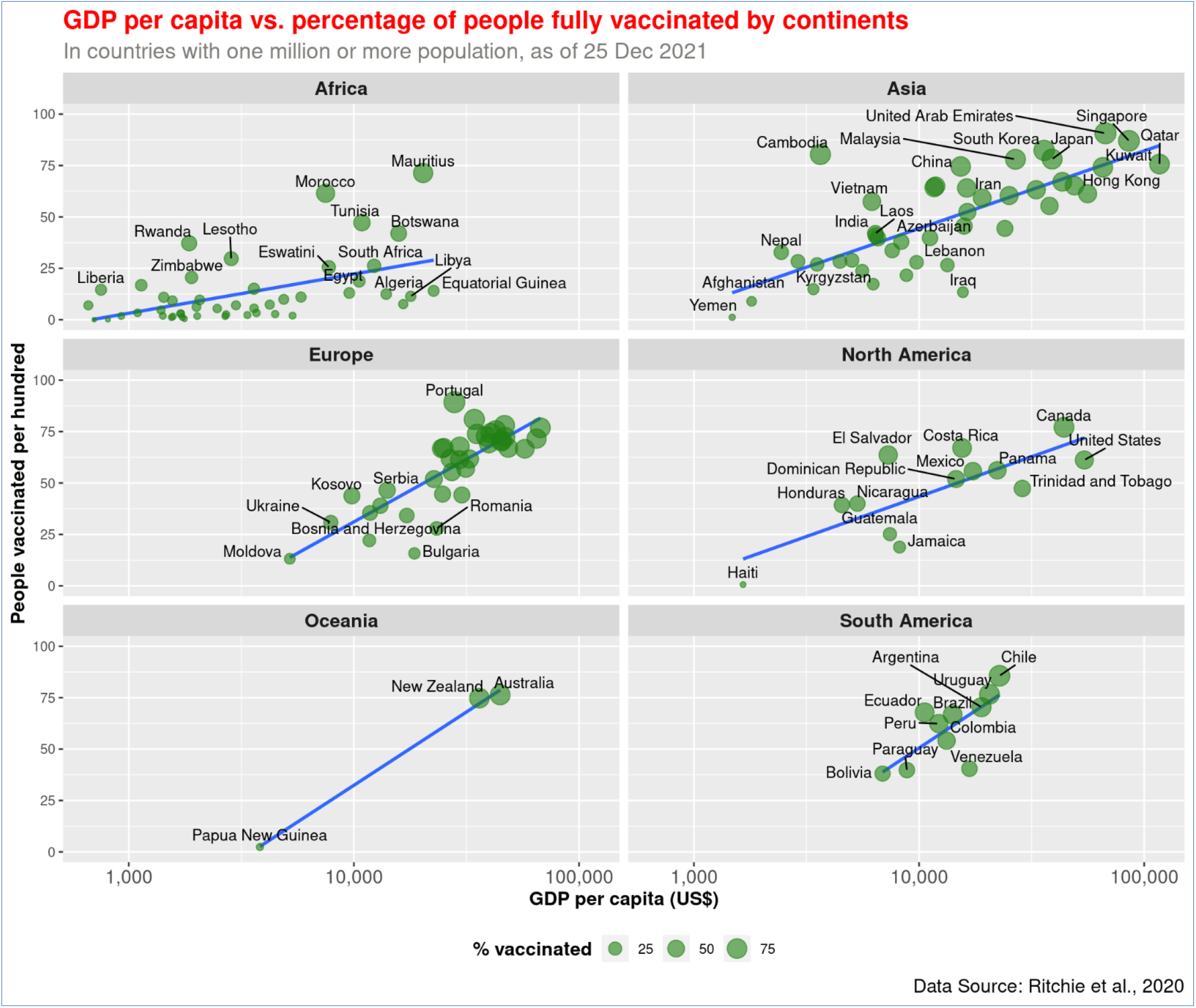
Association of GDP per capita and COVID-19 full vaccination rate across continents. The relationship of both partial and full vaccination rates with GDP is similar in the continents.

The highest level of disparity in GDP per capita and vaccination rate has been observed in Asia. While less-developed Asian countries like Yemen, Afghanistan, and Kyrgyzstan vaccinated less than 20% of their population, more than five countries in the continent achieved over 80% of vaccination. India performed relatively better in immunizing people compared to its wealth, and the country has achieved approximately 60% vaccination. Being a country with modest GDP, Bangladesh follows the trend but has made better progress in recent months. About 53% population of Bangladesh has received at least one dose of COVID-19 vaccination so far. UAE performed better than Kuwait even though their national income level is similar. Yemen performed the worst in Asia.

In Europe, countries with a lower GPS per capita, such as Moldova, Bulgaria, and Bosnia and Herzegovina, obtained 26% or fewer vaccination rates. On the other hand, Portugal, Spain, and Denmark achieved over 80% vaccination, and their GDP per capita is relatively high. Twenty countries in Europe – including the United Kingdom, France, and Germany – attained 63% or more vaccination rates, and their GPP is over US$ 24,000. In Oceania, less developed Papua New Guinea vaccinated about 3% of people while Australia and New Zealand got over 77% vaccination.

Canada (83%) and the United States (US, 62%) performed the best among the North American countries. Countries like Haiti, Jamaica, and Guatemala managed to vaccinate less than 35% of their population in the same continent. In South America, Venezuela appears to make less progress in vaccination compared to its wealth – it managed to have approximately 64% vaccination. The rest of the countries in the continent got vaccination rates proportional to their GPD per capita, and Chile and Argentina from South America obtained over 82% vaccination rates.

The spatial distribution pattern reveals that African nations are least protected against COVID-19 (Figure 6). North American and European nations made the highest level of progress in vaccinating their population. Low-income African countries could not make much progress in the vaccination program during the last few months. There is a significant variation in the vaccination rate among Asian countries (Figure 5). However, the variable vaccination rate in Asia is explainable with the differences in income level.

**Figure 6.**
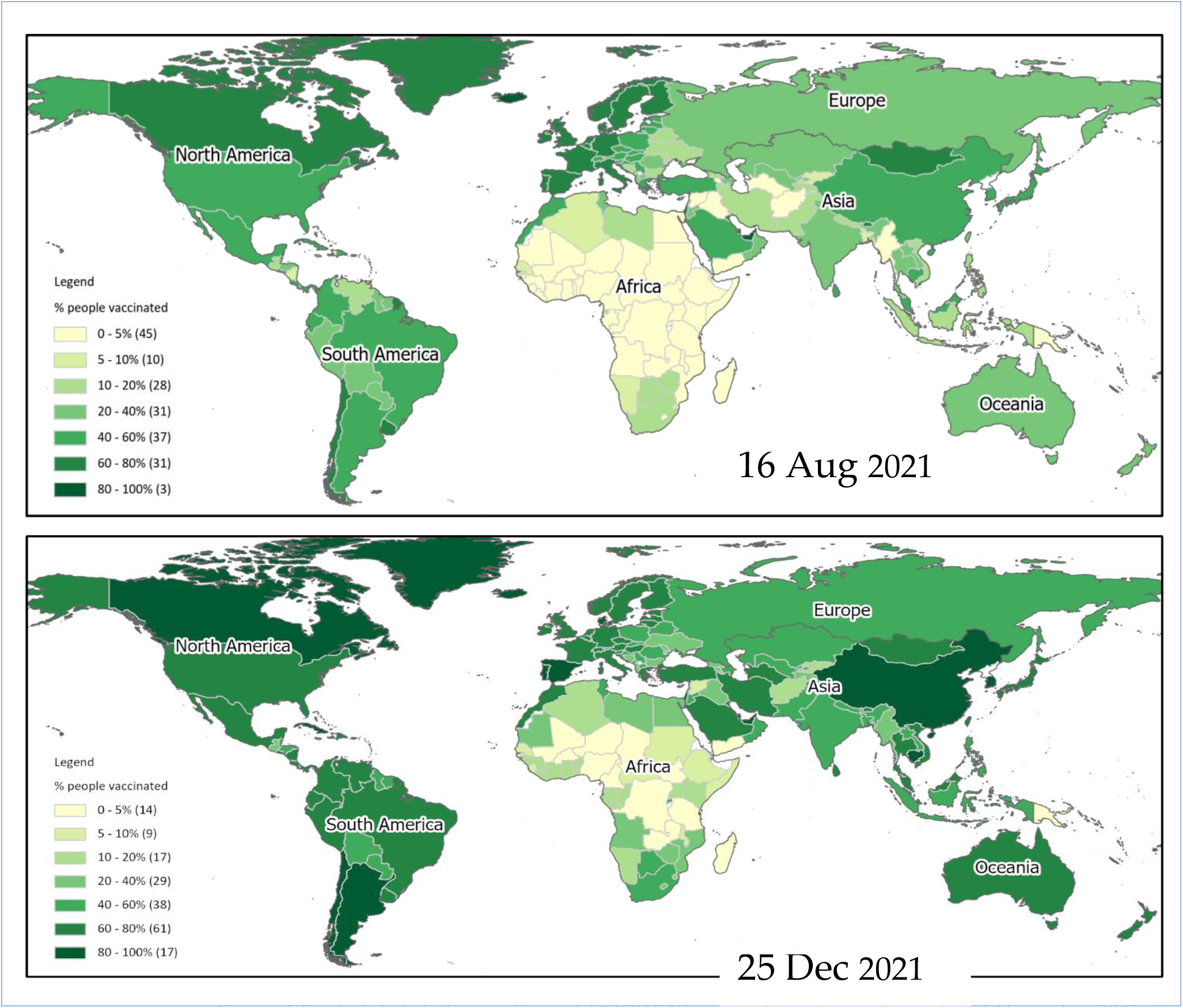
Spatial distribution pattern of COVID-19 vaccination during August and December 2021. Countries in the American and European continent made the highest progress in providing at least one dose of vaccination to their population. Low-income African countries are still lagging behind.

## 4. Discussion

In this current study, we aimed to establish the association between the GDP of nations with COVID-19 vaccination rates. We found that, in general, the wealthier a country is, the higher the vaccination rate. In contrast, the bottom three countries are the Democratic Republic of Congo (DRC), Haiti, and Chad. GPD of these countries is US$ 1,700 or less, and their vaccination rate is lower than 1%. Two of the three countries, DRC and Chad, are in Africa. It has thus been revealed with COVID-19 vaccination progress that humanity is divided into “haves” and “have-nots”. Developed countries secure their population with vaccination, unlike their counterparts in the developing world, where vaccination rates are slower. The important takeaway message is that the COVID-19 vaccine rollout is uneven and that countries in the Global South are not performing as well as their Global North counterparts. Therefore, there are large country-level disparities in being able to achieve herd immunity, and efforts toward attaining global herd immunity are severely hampered. Low- and middle-income countries (LMICs) have always wanted technological and medical advancements (vaccination and drugs inclusive). Regrettable examples include the extensive deaths from HIV/AIDS between 1997 and 2007 in Africa, a disturbing record of 12million deaths that devastated the continent (Nkengasong, Ndembi et al. 2020). While industrialized countries had made the drugs for the disease amply accessible, African countries continued to surrender to the virus. A similar occurrence was observed during the 2009 swine-flu epidemic (Nkengasong, Ndembi et al. 2020). Wealthy countries procured the vaccine for the illness at surplus rates, whereas the economically less endowed countries were found wanting. Unlike the higher-income countries that were able to enforce lockdowns and initiate physical distancing protocols, LMICs have barely been able to do the same, and consequently rearing a blossoming population of more vulnerable individuals. This does not suggest that the richest countries are acting against the poorest ones. Furthermore, the authors do not suggest that the richest countries have to be involved in the poorest countries’ decisions, such as research investment, etc.

Furthermore, LMIC countries have deplorable infrastructures, including bad roads and housing, resulting in an enormous number of difficult-to-reach populations. Despite these, and even though their financial contributions to vaccine development have been limited, the world must carry them along in vaccination protocols towards a common enemy, a no-respecter of the colony or social backgrounds. Developing countries ought to be supported in ensuring access to the COVID-19 vaccine by levelling the power dynamics that perpetuate inequality and fuel injustice (Adebisi, Ekpenyong et al., 2021).

In the past, reports have indicated the inability of African countries to comfortably obtain vaccines due to the high cost. Most, if not all African countries, fall into the category of LMICs (WHO 2020) with other typical African challenges such as poor infrastructure, shortage of health workforce, different religious denominations, and random distribution of population densities (Ophori, Tula et al. 2014), combined with ongoing conflicts and displacements of people. Vaccine research and development are scarcely headed by African countries, which greatly affect their ability to acquire the vaccines they require.

The population of the African continent is estimated at more than 1.2 billion people. With the recent spread of the virus on the continent, more than 1,528,000 people have been infected and more than 36,828 dead (ECDC 2020). This is against the global figure of more than 35,848,000 infected persons and 1,048,000 dead (ECDC 2020). There should be enough supply of vaccines to take care of a good proportion of the continent’s impoverished, sick, and vulnerable populations. Provisions for vaccine storage for subsequent vaccination cases through vaccination programmes are also necessary. These figures reveal the extent to which the urgency for adequate provision of vaccines in Africa can be rated compared with other continents of the world. With Africa constituting 16.7% of the world’s population, the continent ought to compete with other regions of the world for access to the COVID-19 vaccines.

Several HICs who could donate vaccines to Africa are currently under serious strains from the pandemic’s rage, which may have limited the probability of helping developing countries gain access to COVID-19 vaccines. The majority of HICs have made room for vaccines (Edward-Ekpu U 2020) before the call for equitable access to vaccines. The issue of nationalism over universal health coverage (Prabhala and Elder 2020) comes to play as these countries have made early arrangements for the quantities of the vaccine which they would require, a gesture which ultimately tightens the rate of production of the vaccine, which could get to the most vulnerable groups in the least possible time (Shah 2020). Developing countries rely on HICs that manufacture these vaccines to hold back the prevalence of diseases in those countries (Ikilezi, Augusto et al. 2020). Because of the difference in the economic settings of HICs and LMICs, and with the production of vaccines in HICs by pharmaceutical companies (Prabhala and Elder 2020) and research laboratories, the cost of purchasing those vaccines is usually expensive for most poor countries. Nigeria, a typical African country and the most populated, has a high level of under-vaccination due to several challenges, which majorly indicates the high cost of the vaccine (K 2013). Furthermore, in previous years, the vaccination got to the rural communities in Africa late, and even COVID-19 intervention programmes did reach rural communities early (Ogunkola, Adebisi et al. 2020).

With regard to the difficulties faced by potential donor countries of the COVID-19 vaccines, there are indications that countries of the developing world are not being assisted in their bids to assess the vaccines early enough (Edward-Ekpu U 2020, Shah 2020). Most developed countries have placed precedence on meeting their health requirements. Some reports have indicated that the majority of these nations had struck deals to secure vaccines for their citizens (Edward-Ekpu U 2020, Prabhala and Elder 2020). Several of these deals are tied with pharmaceutical companies (Prabhala and Elder 2020). Such countries would no doubt place priority on their needs. Recent reports have indicated the rise in the expenses made by donor nations. Many countries have made deals that have involved massive financial commitments (Edward-Ekpu U 2020) toward responding to COVID-19 and the acquirement of vaccines when ready. This drags the developing countries out of the list of priorities by the donor countries (Edward-Ekpu U 2020).

This current study aimed to establish the association between nations’ GDP with COVID-19 vaccination rates. It has been revealed with COVID-19 vaccination progress that humanity is divided into “haves” and “have-nots”. Developed countries are securing their population with vaccination, unlike the developing countries. The important takeaway message is that the COVID-19 vaccine rollout is uneven and that countries in the Global South are not performing as well as those in the Global North. Therefore, there are large country-level disparities in being able to achieve herd immunity, and efforts toward attaining global herd immunity are severely hampered. One of the biggest challenges that may be observed in the equitable global distribution of COVID-19 vaccines is that there is a disconnect between how some countries agree to do things to show global unity among leaders and the real implementation of the agreement.

A perfect example is the COVAX facility. COVAX is a global initiative established to ensure the speedy and equitable acquisition of COVID-19 vaccines for all nations, irrespective of income level. Low-income countries obtain support from wealthier ones and other donors for vaccine purchases through the Advance Market Commitment (AMC) to make sure that these countries can access the new vaccines simultaneously as the lower-middle, middle, upper-middle, and high-income countries. However, in practice, the scenario is a bit different, which was the fear of the WHO and public health advocates. Internationally, high-income countries have adopted a competitive “fend for yourself” attitude, competing against others for access to supplies and commercial advantage in the COVID-19 vaccines.

Most of the leading vaccines supply was pre-ordered by wealthy nations, even before the safety and efficacy data was made accessible. Therefore, the nationalistic competition for vaccines is a key factor contributing to the challenges faced in the equitable global distribution of the COVID-19 vaccines. These practices are contrary to the global interest and are likely to harm countries and citizens of the Global South. Even in Ghana’s situation, where the country (which became the first country to receive a shipment of the vaccine from the COVAX initiative, but, currently, the supply is just enough for one percent of the country’s population) recently received 600,000 doses of the AstraZeneca/Oxford vaccine, it is seen that this is still too low of a supply to be able to achieve herd immunity. The global pattern observed is that the low-income countries with the least economic and political bargaining power have less access to vaccines.

Foremost, to adopt an attitude, an approach, and actions that reflect global fairness, solidarity, and equity for the expedient vaccine distribution and immunisation campaigns across the globe in order that there is not a discrepancy between words and actions. The WHO director-general, Dr Tedros Adhanom Ghebreyesus, stated it clearly in his opening speech of a WHO executive board meeting: “Not only does this me-first approach leave the world’s poorest and most vulnerable people at risk, it is also self-defeating. Ultimately, these actions will only prolong the pandemic, prolong our pain, the restrictions needed to contain it, and human and economic suffering” (WHO 2021). This recommendation also resonates with the idea that no one is safe until everyone is safe. There is the risk of exacerbating more inequities in COVID-19 infection and mortality rates with some of the current practices.

Furthermore, there is the need for local, national, regional, and international coordination of the vaccination rollout. There is already an established mechanism to actualise this. It is known as the Access to COVID-19 Tools (ACT) Accelerator, a partnership launched by WHO and its partners to support this coordinated and global effort (WHO 2021).

## 5. Conclusions

There is the need for both a top-down and bottom-up approach; that is, a strong commitment, cooperation, and implementation of plans among our scientific, industrial, and political leaders in conjunction with community mobilization at the local levels. The important message driving these recommendations is that, given that the COVID-19 pandemic is impacting a global scale, there is a need for a global response instead of a one-region at a time high-income-countries-before-all others type of response to control the pandemic. The wealthier nations will not be secured without adequately vaccinating the poor ones. Therefore, the global community should take initiatives to speed up the COVID-19 vaccination program in all countries of the world, irrespective of their wealth.

## Data Availability

All data produced are available online at https://ourworldindata.org/

https://ourworldindata.org/

## Supplementary Materials

The following supporting information can be downloaded at: https://rstudio.cloud/project/2771953, R-Studio Project files including R-code and raw data used in the analysis.

## Author Contributions

Conceptualization, P.B., T.A. and K.E.A.; methodology, P.B. and T.A.; software, P.B.; validation, P.B., N.R.Z. and T.A.; formal analysis, P.B.; investigation, P.B.; resources, A.H.M.; data curation, P.B., T.A. and K.E.A.; writing—original draft preparation, P.B. and T.A.; writing—review and editing, A.H.M., N.R.Z., M.K., R.H., A.M. and K.E.A.; visualization, P.B.; supervision, K.E.A.; project administration, T.A. and R.H.; All authors have read and agreed to the published version of the manuscript.

